# Hygiene and attractiveness consequences of Speedos versus swimming shorts: results of a cross-over study among male academics

**DOI:** 10.1101/2025.10.26.25338829

**Authors:** Wouter Graumans, Will J. R. Stone, Jordache Ramjith, Chiara Andolina, Carla Proietti, Sarah Merkling, Ronald van Rij, Merel J. Smit, Heiman Wertheim, Teun Bousema

**Author notes:** **Correspondance to:** Wouter Graumans Department of Medical Microbiology. Radboud university medical center. PO Box 9101, 6500 HB, Nijmegen, the Netherlands. Share joint first authorship (Wouter Graumans, Will J. R. Stone).

## Abstract

Whilst in most northern European countries swimming shorts are the norm, swimming briefs, or Speedos, are compulsory in France and parts of Italy. Hygiene is the main justification for this harsh measure, but empirical data is lacking and consequences for psychological well-being remain unaddressed. This study aimed to empirically assess the hitherto theoretical risk to public pool hygiene posed by swimming shorts, as compared to form fitting swimming briefs (Speedos) that are required swimwear in several southern European countries.

The study design was a cross-over study and photo elicitation, involving international adult male academics and technical staff, of varying age (20-54), BMI (17.8-25.7), and job title. Speedos or swimming shorts were worn for two hours during the normal working day to assess bodily contamination, or as swimwear in natural water bodies to assess environmental contamination. The primary outcome was the number of colony-forming units (CFU) of Gram-positive bacteria (GPB) and Gram-negative bacteria (GNB) on agar plated with 100 µL water collected from i) submersion of the swimming garments in clean tap-water (bodily contamination), or ii) residual water from natural sources (environmental contamination). The main secondary outcomes were self-perceived and externally assessed attractiveness of anonymized photos of the trunk (top of legs to neck) of study participants wearing both swimwear types.

GPB and GNB growth was significantly higher after incubation with tap water rinsed from submersed shorts compared to Speedos (rank-sum, GPB p<0.0400, GNB p=0.0100). Swimming in natural water resulted in considerably higher levels of GNB on Speedos (median 600 CFU/litre, interquartile-range (IQR): 600 to 31400) and shorts (median 33800, IQR: 5200 to 91000), but sample sizes were too low for formal analysis. Participants were significantly less attractive to external judges in Speedos compared to shorts (p<0.0067), but not when judged by themselves (Wilcoxon paired sign rank test: p=0.3394).

We conclude that both GPB and GNB contamination were more abundant in water exposed to swimming shorts, as compared to Speedos. The superior hygiene afforded by scant swimwear comes at the cost of markedly lower externally assessed attractiveness.

## Introduction

Each year, residents of the European Union make over one billion recreational trips (1), with leisure, relaxation and cultural exploration commonly cited as motives for travel. However, some cultural practices encountered during travel can present unexpected sources of discomfort. Public swimming pools, a centerpiece of many family holidays, are notable for their regulatory divergence within Europe, particularly in relation to male swimwear.

Whilst in most northern European countries swimming shorts are the norm, swimming briefs, also known as Speedos, are compulsory attire for access to public pools in France and parts of Italy (**Figure 1**). The justification for these unusually tight regulations is typically framed around hygiene, the suggestion being that looser garments may introduce external contaminants into the pool and its environment (2). However, to the best of our knowledge, these claims have never been substantiated by empirical data.

**Figure 1.**
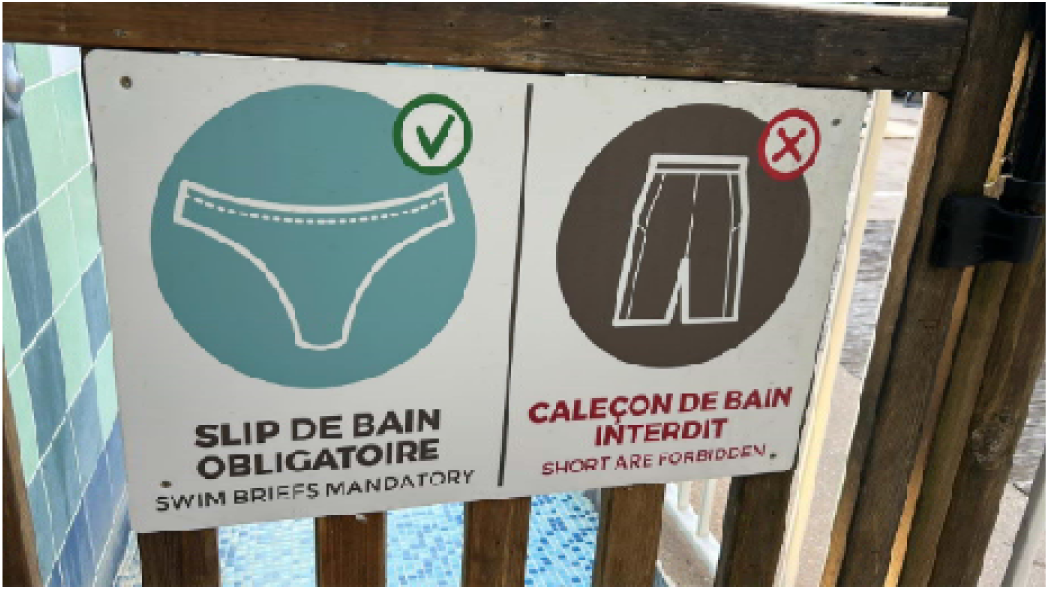
Common signage photographed at French swimming pool.

The lack of evidence supporting these regulations, or lack of transparency if such research has been conducted, is noteworthy considering their potential impact on the self-esteem and psychological well-being of unsuspecting travelers. For visitors without foreknowledge of such policies, hurried purchases may add a not inconsiderable financial burden to the already high cost of holidaying. Those unaccustomed to such policies may also suffer considerable discomfort or embarrassment, particularly in the context of rising levels of body image anxiety (3). Given the large number of citizens affected and with the risk of further fueling tensions between northern and southern European countries at a time when other geopolitical issues demand unity, we aimed to provide definitive evidence on the hygiene of differing male swimming outfits. Suspecting alternative motives for the southern European preference for swimming briefs, we also investigated the attractiveness of male members of academia to a French and Italian panel of observers, directly comparing briefs and shorts. Needless to say, our findings are highly revealing.

## Methods

As necessary preparatory work for our main trial we performed an ownership survey among our Dutch participants, indicating that whilst swimming shorts were abundantly pre-owned, none had - or felt comfortable admitting to the possession of - Speedos. We thus provided the form-fitting swimwear, selecting a design that came highly recommended by the self-reporting public; customers gave the pair a mean of 4.6 out of 5 stars based on 1142 reviews (Decathlon, size NL50/FR44).

Perhaps unremarkably, once assured that they would be receiving free swimwear, an international group of male academics (n=21) provided informed consent to participate in a complex cross-over study to investigate the relative contamination risk posed by both swimming shorts and Speedos. In this main trial, participants were asked to wear Speedos or swimming shorts underneath normal clothes for exactly 2 hours, to test the propensity of different styles of swimming outfit to accumulate pathogenic bacteria after prolonged use. Before beginning their experiments, participants were provided a computer-generated random order in which to test the garments and were instructed to conduct their experiments on back-to-back days, showering an unspecified but uniform time before donning their swimwear; defecation during this time-period was strictly forbidden.

Prior to each round of sampling, we asked our participants to wash both their shorts and Speedos according to manufacturer instructions with a standard detergent. Academic status (technical staff, PhD student, early career researcher, senior researcher [assistant, associate or full professor]) was recorded as a potential co-variate in our analysis, aware as we are that standards of hygiene may be associated with stress, and quality of washing machine may correlate with salary.

After the test garments had been washed, and post-wash bacterial contamination confirmed to be negligible (**Table S1**), participants wore their swimwear for exactly 2 hours. They were then invited to disrobe, carefully wash their hands (>20 seconds with soap), immerse their outfits in 1000mL of tap water, and squeeze them dry. This was repeated four times. Acknowledging a potential role for variation in fabric quantity (up to 5700 cm^2^ for shorts, 4-fold the size of the Speedo at 1440 cm^2^), the water volume extracted was recorded, and 50mL removed and stored at 4°C for a maximum of 24 hours. Water samples (100µL) were plated on MacConkey agar (Thermo Fisher Scientific, without salt, PO0148A) using a Drigalski spatula to determine the number of colony-forming units (CFU) after 24 hours incubation at 35°C. For lab members for whom high levels of contaminants were suspected (e.g. students close to their thesis hand in date, or early career researchers approaching the end of yet another fixed term contract), serial dilutions were made prior to plating. Colonies were counted, categorized Gram-positive bacteria (GPB) or Gram-negative bacteria (GNB), and presented as total CFU per liter.

In our second set of experiments, we aimed to address the concerns of Speedo-adherent public pool-owners that swimming shorts are more likely to be worn in public spaces, harbor contaminants from less controlled swimming environments and thus carry a higher risk of contamination from external environmental sources. To assess the impact of swimwear choice on bacterial accumulation after swimming in non-chlorinated water, we asked a sub-set of participants to swim in open water in both briefs and shorts. Participants were allowed to define the location and duration of swimming *ad libitum* as long as it was identical for both outfits and the random allocation of outfit order was observed. All other methodological rules associated with the swim and sample collection were regulated rigidly.

Adolescent pool-goers are also known to wear their (branded) boxershorts underneath swimming shorts, potentially increasing contaminant risk to pool waters with this use as outerwear. A sub-set of participants were therefore asked to wear boxershorts for two days underneath their swimming shorts prior to immersing both the boxershorts and the swimming shorts in 1000mL of water, as described above.

Lastly, we examined alternative motives for enforcing Speedo use in public pools. We hypothesized that southern Europeans have a preference for the appearance of male torsos in Speedos and/or an aversion to the same torsos in swimming shorts. To test this hypothesis, we asked participants from our main pool of participants to take pictures of their torso (i.e. their trunk – groin, midriff, and chest; photographically decapitated and thereby anonymized) in both Speedos and shorts. Pictures were turned to a grayscale to avoid the unnecessary distraction of color and pattern (often rather vivid on swimming shorts, and bland on Speedos), and to enhance focus on form. French and Italian academics of all genders were asked to judge the attractiveness of the photographed outfits. This was done using two approaches. First, pairs of pictures were shown to the panel of observers, who were asked to select the most attractive outfit for each participant. In addition, participants and observers were asked to rank attractiveness on a scale of 1-10, with 10 being the most attractive. For both approaches, sentinel pictures were included in the form of similarly anonymized pictures of notable individuals in a variety of swimming outfits, including actors Arnold Schwarzenegger, Steve McFadden and Leonardo DiCaprio, swimmer Michael Phelps, comedian Rowan Atkinson, and - King Charles III. These pictures were included to confirm that panel members were humans and submitted honest scores; accurately grouping sentinel pictures in pre-defined bins of attractive and less attractive was taken to indicate that the experiment was assessed trustworthy. We also asked participants to rank their own physical attractiveness (based on (4)). The panel of observants consisted of French academics and Italian academics. Since academics are a highly mobile populations we cannot rule out that swimwear preferences may develop during residency among swimming shorts-favoring populations, current country of residency and time since leaving areas that favour more revealing swimwear were included as interaction terms in our analyses.

### Statistical methods

Analyses were conducted in R, version 3.1.12 (Team RC, 2019). The sample size for the cross-over study was defined based on paired comparisons. With 20 paired observations, we estimated we would have 91% power to detect a statistically significant difference at an alpha of 0.05 if ≥80% of paired observations showed higher CFU for swimming shorts; statistical power increased to and 99.7% if ≥90% of paired observations showed higher CFU for shorts. Whilst the nature of the data prompted us to explore pooled analyses, we opted to include individual observations. The number of CFU was compared between outfits with Wilcoxon paired sing-rank test. Spearman rho correlation tests were used to access correlation between self- and panel assessed attractiveness. For the attractiveness analyses, shrinkage methods were explored to investigate which predictors (age, position, self-rating, country of rater and type of swimwear) are predictive of the external raters ratings. We then ran a mixed effects linear model with all the variables (mentioned above) as fixed factors and random intercepts (using glmmLasso package in R) for individuals to account for intra-individual correlation.

## Results

A total of 21 participants took part in the cross-over study on self-derived bacteria accumulation in different swimwear types. While new Speedos were provided by the study coordinators and were thus uniformly embarrassing in size and shape, swimming shorts were participant-owned and varied significantly, the largest being 5700cm^2^ and the oldest being a venerable 12 years of age. Sampling was done in the first half of July 2025, a period of unusually high temperatures in the Netherlands (min. 19.5 - max. 35.5°C), marking the first official heatwave of the year. The median body mass index (BMI, (5)) of participants was 22.6 (IQR: 20.8 to 24.4); 14% of participants were in the age range from 20-30 years, 29% between 30-40, and 57% 40 years or older; 33% was employed as technical staff, 10% PhD student, 19% early career researcher, and 38% had a senior academic status (assistant, associate or full professor). Activities during the time-window when the primary swimwear experiments were conducted ranged from crispr-cas gene editing and advanced statistical modeling (early career researchers), to sending emails and drinking coffee (senior academics). Though activities concurrent with swimwear wearing varied widely, with some conceivably influencing the gut microbiome and more demanding pursuits potentially increasing the risk of stress-induced flatulence (6, 7), we assumed our cross-over design accounted for these factors, and none were judged to merit exclusion from the study.

During sample preparation, it was noted that considerably more water was retained in shorts (median 150mL, IQR; 100 to 210) than in Speedos (median 65mL, IQR; 26 to 75) (p = <0.0001). After plating water samples on agar plates and incubating at 35°C for 24 hours, a large variation in bacterial colony formation from GPB was observed (range 700 - 181700 CFU). Expressing GPB growth as CFU per liter of water, we consistently observed higher growth after incubation with water from swimming shorts compared to Speedos in our paired analysis (rank-sum, p<0.0400) (**Figure 2A**). Although GNB growth was markedly lower for both outfits (**Figure 2B**, p<0.0001), higher counts were again consistent for swimming shorts (median 0 CFU/L versus 100 CFU/L; p= 0.01). There was no evidence that median CFU was associated with career/academic status.

**Figure 2.**
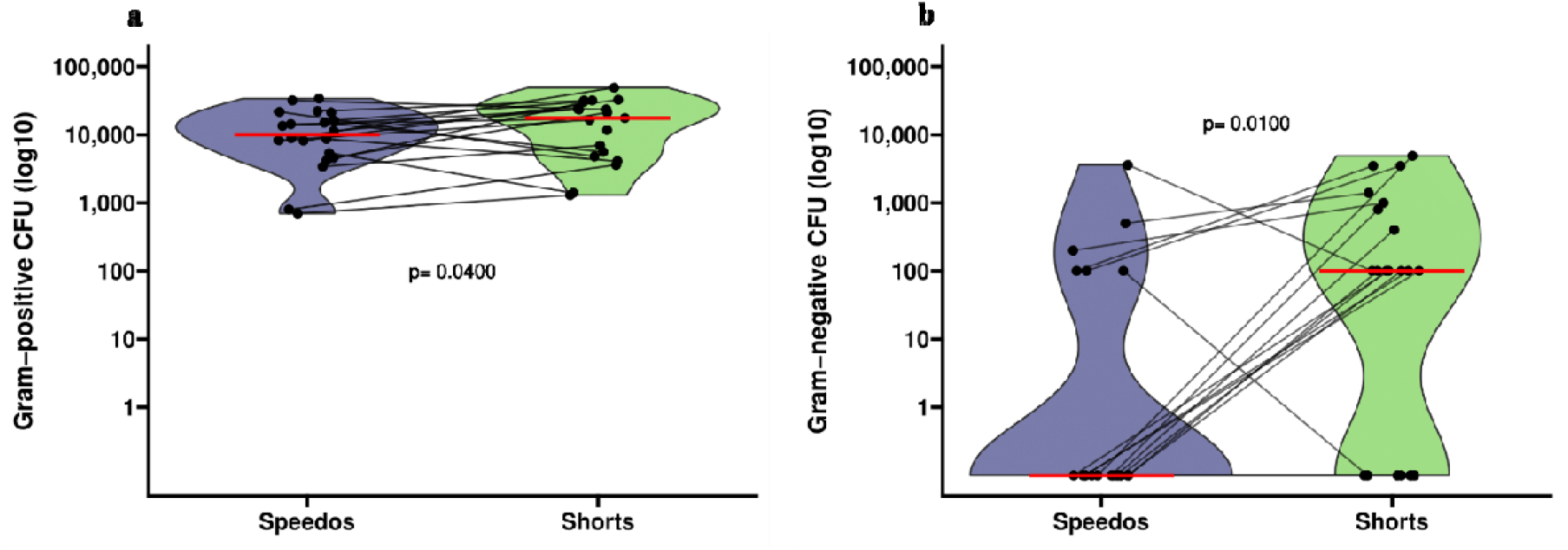
Bacterial load of different types of swimming outfits. Violin plots present findings from samples of 21 participants who wore a standardized Speedo (purple) or self-owned swimming shorts (green) for two hours. The Y-axis presents the number of Colony Forming Units (CFU), expressed per liter of sample to account for differences in retained water volume (p =<0.0001) between shorts (median 150mL, IQR; 100 to 210) and Speedos (median 65mL, IQR; 26 to 75), for Gram Positive Bacteria (GPB; panel A) or Gram Negative Bacteria (GNB, Panel B). P-values are for Wilcoxon signed-rank tests. Median CFU for GPB were 10150 CFU/L (interquartile range; IQR 5200 - 17475) for Speedos and 22350 CFU/L (IQR 5400 - 31650) for shorts; (Panel A; p<0.0400). Median CFU for GNB were 0 CFU/L (IQR 0 - 100) for Speedos and 100 CFU (IQR 0 - 852) for shorts; (Panel B; p= 0.01).

To assess the impact of variable water quality on the accumulation of environmental bacteria on swimming outfits, water from paired samples of shorts and Speedos were collected after swimming experiments in local waterbodies by a sub-set of more adventurous participants (n=5). In at least two of these experiments, local authorities had issued water-quality warnings to recreational swimmers, likely the result of a drought in Europe which led to widespread decreases in water levels and heightened pollutants. We considered this an exciting opportunity to explore the full potential of swimming outfits as bacterial carriers, and a sign of our participants dedication to answering one of mankind’s great questions. Sample collection in the wild-swimming experiments was rather eventful. In one unfortunate occurrence, a participant’s clothes were stolen by a person not involved in the trial or known to the participant, leaving the participant in a slightly embarrassing outfit in public negotiating return of the clothes with the stealer. In a second event, a participant left his Speedo to dry on a rock while swimming in his shorts. A dog (*Canis lupus familiaris*) briefly urinated on the Speedo. Whilst urine is a sterile fluid, this sample was excluded from analysis and the apologies from the dog owner were accepted. Swimming in natural water resulted in considerably higher levels of GNB on Speedos (median 600 CFU/L, IQR; 600 to 31400) and shorts (median 33800, IQR; 5200 to 91000) compared to those observed after wearing outfits underneath clothes (Speedos: median 0, IQR; 0 to 100; Shorts: median 100; IQR 0 to 850).

In a final, possibly misjudged experiment to assess bacterial accumulation due to misuse of swimming shorts as outerwear (i.e. with underwear), in which against all hygiene standards, boxershorts were worn for two consecutive days underneath swimming shorts, we observed unsurprising in number of GNB (median 20400 CFU/L, IQR; 10500 to 38200).

In a uniquely open-minded international collaboration, anonymized pictures of 11 participants were shared with 11 French academics (7 female, 3 male, 1 non-binary) and 24 Italian academics (20 female, 4 male, 0 non-binary). After reassuring ourselves votes for sentinel pictures gave no reason to suspect that panel members did not appropriately value the scientific rigor or importance of this study, there was overwhelming evidence that our participants were less attractive in Speedos compared to shorts by both the French (p=0.0049) and the Italian panel members (p=0.0067) (**Figure 3A**). The general inferiority of Speedos in terms of physical attractiveness was only partially corroborated by a self-assessment of attractiveness. Whilst the Southern-European panel had a clear preference, there was a tendency for participants scoring their own attractiveness slightly higher in shorts but no significant difference compared with speedos (Wilcoxon paired sign rank test: p=0.3394). Participants also strikingly overrated themselves, both in shorts and speedos (French vs self-rated Speedos p=0.02338; French vs self-rated shorts p=0.0234; Italians vs self-rated Speedos p=0.02938; Italians vs self-rated shorts p=0.0232). Of note, several senior academics scored their own attractiveness markedly higher than our panel of observers, the weak positive correlation (spearman rho = 0.22) is potentially reflecting an unrealistic positive perception of performance of associate and full professors that has also been reported for more academic qualities (8) (**Figure 3B**).

**Figure 3.**
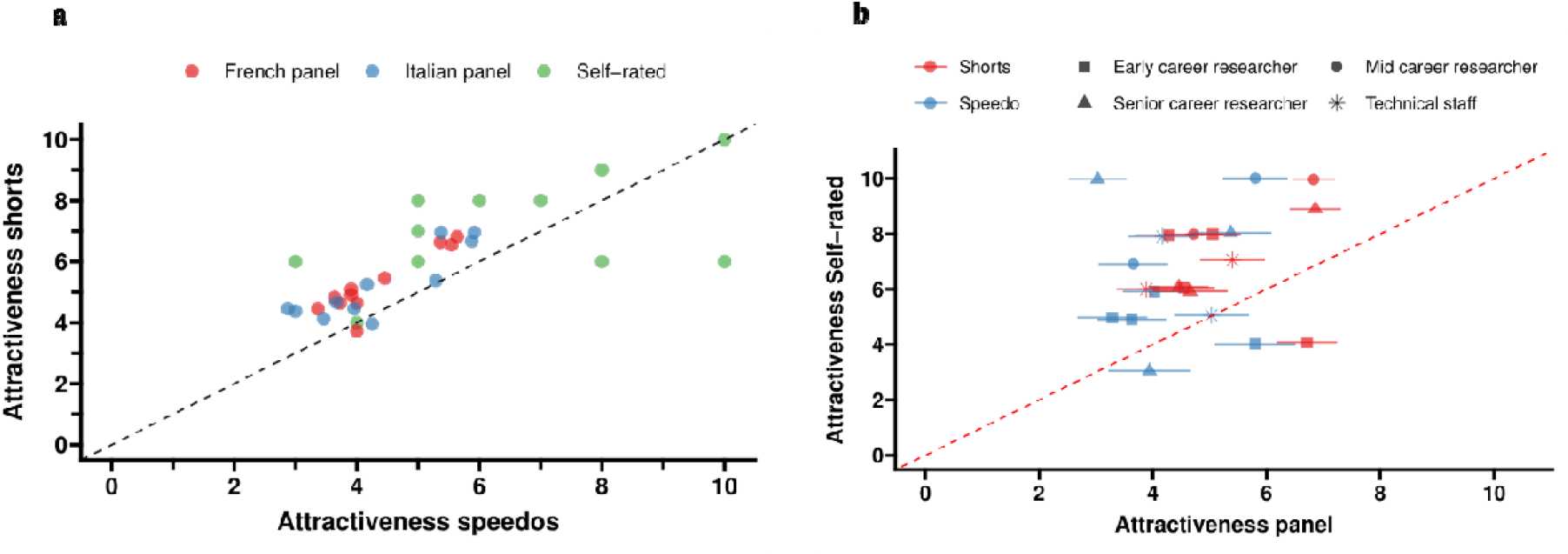
Attractiveness of different types of swimming outfits. The correlation plot presents attractiveness on a scale of 1-10 in Speedos (X-axis) and swimming shorts (Y-axis), as scored by study participants themselves (in green; n=11), or French (blue, n=11) and Italian (red, n=24) panelists. The dotted line reflects perfect agreement between scores; actual scores are overwhelmingly higher for shorts (panel A). Correlation plot presents attractiveness of academic panel (mean and 95% CI) and self-rated attractiveness. Symbols indicate academic status. The dotted line again reflects perfect agreement; panel members gave lower scores in attractiveness than volunteers gave themselves (panel B).

## Discussion

We aimed to provide definitive evidence on the bacterial contamination risks to public swimming facilities posed by different male swimwear options. We also investigated the potential for ulterior motives – namely, aesthetic preference - among individuals residing in locales where speedo mandates are enforced, questioning whether hygiene is truly the driving factor behind these regulations.

Much to our surprise we find strong evidence that both GPB and GNB are more abundant in water exposed to swimming shorts, as compared to swimming briefs (or Speedos). The Netherlands Pool Directive (Zwembadrichtlijn) considers a safe threshold for bacterial contamination of up to 1 CFU/100mL of *enterococci* (GPB) and/or *pseudomonas* (GNB). To contextualize this, consider a standard sized recreational pool of 12m by 6m (1.5 m depth), containing approximately 105 thousand liters of water. Based on the average GNB bacterial load observed in our study, it would take approximately 1044 pairs of Speedos or 19 swimming shorts to raise contamination levels over safe thresholds. However, perhaps as a warning against university away-days, a mere 47 speedos or 1 pair of shorts from our most contaminant prone participant (senior academic) would suffice to convert the pool into a potential biohazard. Strict regulation and diligent chlorination are of course the best form of protection against unsanitary academics. However, though chlorination is still the mainstay of prevention, it is not a cure-all and is reliant on correct pool maintenance. We confirmed that the bacterial load released from swimwear is potentially controlled by chemicals added to the pool water but also observed that this did not annul contamination (**Table S2**).

There are several possible reasons why Speedos may associate with lower bacterial loads. It is possible that contaminant release from the gastrointestinal tract is lower in Speedos due to their elasticity exerting external pressure on the gluteal muscles, thereby reducing contact between the rectum and the fabric. Another hypothesis associated with the loose-fitting nature of swimming shorts, is that when swimming, bacterial accumulation might be impacted by fluid dynamics. Surprisingly, the impact of pool hydrodynamic drag on fecal bacterial shedding is grossly unexplored and to the best of our knowledge, no studies have ever examined fluid dynamics inside different types of swimwear. Whilst such studies would cast an important light on the risks posed by loose swimwear to public pools and their occupants, it is unlikely to have affected our main findings where swimwear was worn under dry conditions. Instead, we consider it plausible that, like many Northern Europeans, microbes may be inherently averse to Speedos. Swimming shorts and briefs are typically constructed from materials suited to their respective purposes. Briefs prioritise speed and performance, utilising material blends including nylon, elastane and spandex for a snug fit and reduced drag. Swimming shorts, on the other hand, are designed for comfort and versatility, often using materials like polyester, nylon, or blends of these with natural fibers. In the current trial, our Speedos were made from a fabric blend of 18% elastane with 82% polyester while the shorts (pre-owned by our study participants) were a mix of synthetic materials (e.g. polyamide, polyester) and natural material (cotton, linen). Fabric surface properties like hydrophobicity and hygroscopicity influence bacterial colonization and could be modified to reduce biofilm formation (9).

One of our study limitations was we were unable to sufficiently account for what constitutes ‘normal use’ for our swimming outfits. While we assert that our paired experiments accurately represent the *inherent* risks posed by (previously) clean garments to public pool contamination, we acknowledge that our study design does not reflect real world use, which may be heavily affected by more frequent and unconventional use of swimming shorts as outerwear. This is arguably the most intuitive reason to oppose swimming shorts in public pools. Our sparse self-reported data from participants working mostly in biosafety level 2 and 3 laboratories, suggests that prolonged wearing of Speedos as casual wear is indeed unlikely. We were struck by the low attractiveness scores, as provided by external panel members. The authors of this report, most of whom were also participants in the main trial and attractiveness studies, see no reason to consider themselves less attractive than the general public and – impartially – consider it probable that the external scoring, being as it was performed exclusively by academics (a profession tending toward excessive criticism) may have been unduly harsh.

In summary, we present moderate evidence that the requirement for use of swimming briefs, as opposed to more conservative shorts, is justified by the oft cited but hitherto unevidenced claim that shorts pose a greater contamination risk to public pools. Future studies should seek to clarify whether design innovations (i.e. anti-bacterial fabrics) can mitigate the contamination risks of looser swimwear, without compromising comfort or self-respect.

## Data Availability

The datasets used in this study are available upon reasonable request.

## Acknowledgements

We would like to acknowledge Matthew McCall, Myriam Hanskamp-Vermeeren, Mariëlle Wendling and Hester Vogelaar for providing microbiological expertise and support. The authors are immensely grateful to the participants who contributed to this work. Those that have expressed a specific wish to be acknowledged as participants here follow: Nick Proellochs, Matthew McCall, Taco Kooij, Jakko van Ingen, Mike Mientjes, Simon Koele, Gijs Overheul, Twan Klaassen, Felix Hol, Pascal Miesen, Óscar Lezcano, Paul Rutten, Tom Otter, Jeroen Bok, and Joep van Erp. Special thanks to Swimschool M. Laport for providing access to public pools in Ede and Renkum, The Netherlands□□.

## Availability of data and materials

The datasets used in this study are available upon reasonable request. The Speedos used in this project are available to whoever is willing to pay for postage.

## CRediT authorship contribution statement

Conceptualization by Teun Bousema; data collection by Wouter Graumans, Chiara Andolina Carla Proietti and Ronald van Rij; statistical analysis was performed by Teun Bousema, Jordache Ramjith and Will Stone; all authors wrote – reviewed & edited the initial draft; all listed authors meet authorship criteria, endorsed the final version of the manuscript.

## Ethics approval and consent to participate

The study protocol, consent form and questionnaire were submitted to the Medical Ethics Review Committee (METC) Oost-Nederland (dossier 2024-17621). According to the Medical Ethics Review Committee (METC) Oost-Nederland, this study (dossier 2024-17621) does not fall under the scope of the Dutch Medical Research Involving Human Subjects Act (WMO) and therefore did not require formal ethical approval. All participants provided informed consent.

## Supplementary material

**Table S1:** CFU/L Gram-positive and -negative bacteria after sampling of washed Speedos and trunks according to manufacturer instructions with a standard detergent.

| Sample ID<br>(Speedo/trunk) | Gram-positive bacteria (CFU/L) | Gram-negative bacteria (CFU/L) |
| --- | --- | --- |
| Control 1 (Speedo) | 1200 | 0 |
| Control 2 (Speedo) | 1600 | 0 |
| Control 1 (trunk) | 1100 | 0 |
| Control 2 (trunk) | 1700 | 0 |
| Control 3 (trunk) | 0 | 0 |
| Control 4 (trunk) | 1500 | 0 |

**Table S2:**
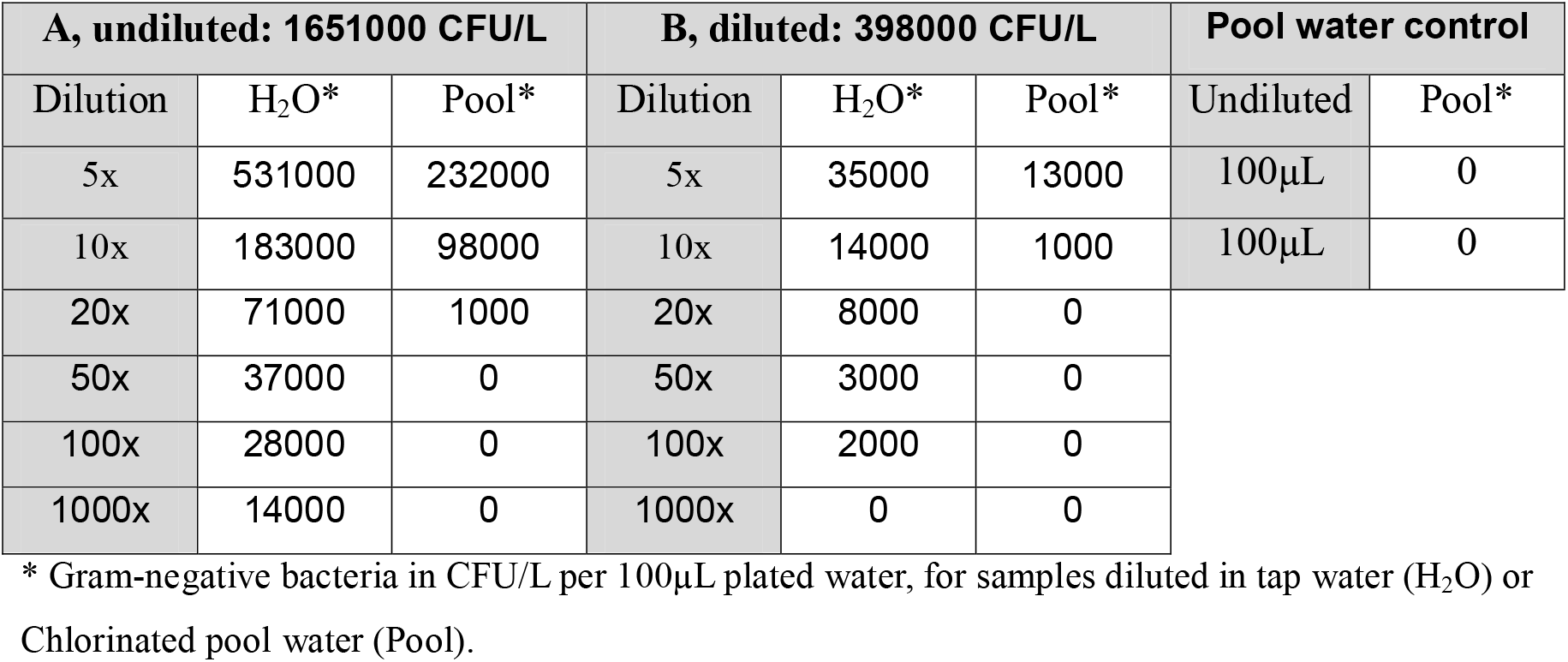
Effect of chlorinated pool water on a highly contaminated pair of garments, for an undiluted (**A**) and a diluted sample (**B**).

